# Vaccination scenario-based study on seasonal influenza in the Republic of Korea

**DOI:** 10.1101/2025.03.27.25324818

**Authors:** Vijay Pal Bajiya, Jongmin Lee, Eunok Jung

## Abstract

**Background:** Influenza is a major global public health issue, particularly affecting older adults aged 65 years and above. Vaccination is the most effective strategy for preventing the spread of seasonal influenza and is recommended by the World Health Organization for high-risk groups. However, the prioritization and timing of vaccination across different risk groups are crucial factors affecting the success of vaccination programs.

**Method:** In this study, we employed an age-structured SEIAHR model using seasonal influenza data from the Health Insurance Review and Assessment Service, and the influenza vaccination program in South Korea, as part of the Korean National Immunization Program. The transmission coefficients for various age groups and seasonality factors were estimated.

**Results:** Our analysis reveals that under the baseline vaccination strategy, the highest infection rates occur in the G1 age group (children aged 0 to 14 years) and the early vaccination of the G4 group (individuals aged 65 years and older) is crucial for reducing severe outcomes. Specifically, scenarios prioritizing early vaccination for the elderly resulted in a reduction in cumulative cases by approximately 34% and a 45–60% decrease in peak infection levels compared with the baseline.

**Conclusion:** These findings underscore that the targeted and timely vaccination of high-risk populations not only mitigates the overall epidemic burden but also shortens the period during which hospitalization thresholds are exceeded. Our study demonstrates that age-specific vaccination strategies, particularly those that accelerate vaccination in elderly individuals, are essential for minimizing the public health impact of seasonal influenza.

## Introduction

Influenza is a major public health challenge across the globe, with a particularly significant impact on older adults aged 65 years and older. Every year, the virus affects 5–10% of the adult population and 20–30% of children worldwide [1]. It is responsible for causing between three and five million cases of severe illness, along with an estimated 290,000 to 650,000 deaths annually [2]. In the Republic of Korea, influenza causes approximately 400,000 outpatient visits and 7,000 hospitalizations annually [3, 4]. Older adults in the country face a disproportionately high risk, with a mortality rate of 46.98 per 100,000, which is far greater than the general mortality rate of the population of 5.97 per 100,000 [3]. This difference underscores the elevated vulnerability of elderly individuals to influenza complications, highlighting the urgent need for targeted prevention and treatment strategies.

As a part of the Korean National Immunization Program (KNIP), the influenza vaccination program in the Republic of Korea primarily targets specific high-risk groups to prevent seasonal influenza. These groups include children under the age of 13 years, adults aged 65 years and older, and pregnant women, all of whom are considered particularly vulnerable to the flu [5]. The aim of the program is to ensure that these high-risk individuals receive a vaccine to minimize the impact of the flu season.

Regarding high-risk populations for seasonal influenza, the focus is on children younger than 5 years, adults aged 50 years and older, pregnant women, and individuals suffering from chronic health conditions [5]. These groups are more likely to experience severe complications from the flu and their vaccination is a priority to protect public health. In the current influenza season, national vaccination coverage is approximately 40 Mathematical modeling is a valuable tool for studying transmission dynamics and evaluating intervention strategies, with many researchers analyzing seasonal influenza epidemics [6]. Given potential pandemic scenarios and challenges in vaccine production [7], early data on age-specific incidence and mortality can help prioritize vaccine distribution and optimize disease burden reduction. Chowell et al. [8] investigated an age-structured model in which the transmission rate for each age group was assumed to be influenced by contact rates. Age-specific risk of illness and hospitalization rates were estimated based on incidence data. Their findings indicated that an adaptive strategy targeting individuals aged 6–59 years was the most effective approach to reduce hospitalizations and deaths. Ratti et al. [9] investigated an SEIR epidemic model to assess optimal vaccination strategies for seasonal influenza in a nursing home with a long-term care environment. They incorporated contact matrices based on surveys of both healthcare workers and residents. Their findings indicate that prioritizing the vaccination of residents, rather than healthcare workers, may still be the most effective preventive approach. Diana and Gergely [10] explored an age-structured compartmental model with vaccination status to assess the effects of age-specific vaccination scheduling during an influenza outbreak. Their findings highlighted that the timing of vaccination by age group can significantly influence the outcome of an epidemic. Knipl and Roast [11] explored an age-structured compartmental model to assess the impact of age-specific vaccination during a pandemic and found that targeted scheduling can significantly affect the epidemic outcome, with delays worsening the infection rate. Mylius et al. [12] developed a model for the Asian flu, recommending prioritizing high-risk individuals when a vaccine becomes available and targeting high-transmission groups if vaccination starts early. Mantel et al. [13] evaluated seasonal flu vaccinations in Belarus, Morocco, and Thailand, emphasizing that healthcare workers are a key target group because of their role in influencing acceptance and ensuring vaccine delivery. McMorrow et al. [14] analyzed influenza data from South Africa to prioritize vaccination risk groups. Tsuzuki et al. [15] found that vaccinating children under 15 years of age has a greater epidemiological impact than targeting the elderly, although Japan’s current program focuses on older individuals. Other studies have emphasized the importance of age-specific intervention strategies and examined the distribution of vaccines across different age groups, assuming pre-seasonal vaccination [12, 16, 17]. In contrast, herein, we focused on prioritizing vaccination across various groups based on the timing of the schedule.

We employed an age-structured SEIHAR model with three distinct vaccination compartments to analyze the dynamics and vaccination strategies for seasonal influenza. To incorporate the age-specific vaccination strategy outlined in the current KNIP, we categorized individuals into four age groups. We examined the variations in influenza incidence rates and influence of distinct contact patterns among age cohorts by estimating the transmission coefficients for these groups using influenza case data from the Health Insurance Review and Assessment Service (HIRA) database. Moreover, we explored tailored vaccination strategies to assess their effectiveness in controlling the spread of influenza. This comprehensive approach enabled us to develop alternative vaccination strategies to the current approach in Korea, aiming to reduce influenza cases and evaluate the impact of targeted efforts on disease incidence across diverse demographic segments.

## Materials and methods

### Data source

We utilized data on influenza cases from August of 2023 to March of 2024, sourced from the HIRA [18, 19] and World Health Organization (WHO) Flunet [20], to develop an age-structured model aimed at examining the transmission dynamics of seasonal influenza in the Republic of Korea. We used vaccination data from the annual report of the KDCA [21, 22], which provides weekly vaccination numbers for high-risk groups.

According to the KNIP for the 2023 to 2024 season, vaccination coverage was higher in certain groups. Approximately 82% of adults aged 65 years and older, approximately 70% of children under 13 years, and 133,735 out of 265,262 eligible pregnant women were vaccinated [21]. Vaccination coverage was assumed to remain consistent across various vaccination scenarios. We composed a contact matrix *C*_*jk*_ of four age groups from the POLYMOD study [23] by considering the population size in each age group. The details of the contact matrix are provided in the Supplementary Information (see S1 File).

### Mathematical Model

Considering the contact patterns, incidence rates, and current vaccination strategy, the total population was categorized into four distinct age groups: Group 1 (G1) for individuals aged 1 to 14 years, Group 2 (G2) for those aged 15 to 49 years, Group 3 (G3) for those aged 50 to 64 years, and Group 4 (G4) for the elderly population aged 65 years and older. In this study, individuals were categorized into different compartments based on their health status, with the subscript *j* representing specific age groups (*j* = 1, 2, 3, 4). The groups include susceptible individuals *S*_*j*_, vaccinated individuals (ineffectively vaccinated *U*_*j*_, effectively vaccinated *V*_*j*_, and those protected by the vaccine *P*_*j*_), exposed individuals *E*_*j*_, infected individuals (asymptomatic *A*_*j*_, symptomatic *I*_*j*_, and hospitalized *H*_*j*_ ), recovered individuals *R*_*j*_, and deceased individuals *D*_*j*_. The total population is defined as 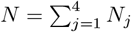,where *N*_*j*_ = *S*_*j*_ + *U*_*j*_ + *V*_*j*_ + *P*_*j*_ + *E*_*j*_ + *A*_*j*_ + *I*_*j*_ + *H*_*j*_ + *R*_*j*_ is the total population of *j*^*th*^ age group.

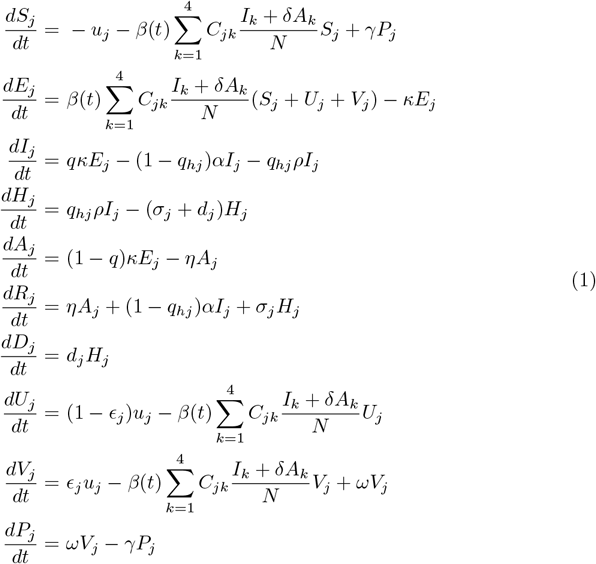

In the force of infection term, *C*_*jk*_ represents the contact rate at which individuals in age group *j* come into contact with individuals in age group *k, δ* is the relative infectiousness of asymptomatic individuals compared with symptomatic individuals, and 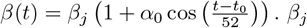 represents the transmission coefficient for the *j*^*th*^ age group, *t*_0_ denotes the seasonality phase, and *α*_0_ denotes the seasonality amplitude of influenza transmission.

The average duration of the latent period and delay before hospitalization are assumed to be represented by 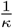 and 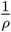,respectively. The proportion of infected individuals who exhibit influenza symptoms is denoted as *p*. Additionally, it is assumed that asymptomatic infected individuals recover at a rate *η* without requiring hospital care. Furthermore, only *q*_*hj*_, namely the age-specific proportion of symptomatic infected individuals, required hospitalization because of severe symptoms, whereas the remaining proportion recovered at a rate *α* without any hospital care. This model also incorporates age-specific recovery rates for hospitalized individuals *σ*_*j*_ and mortality rates (*d*_*j*_). Currently, according to the KNIP, the authorities provide seasonal influenza vaccinations for specific high-risk groups, including children under 13 years of age, adults over 65 years old, and pregnant women. Consequently, the age-specific vaccination rate *u*_*j*_ is considered. However, it is assumed that only *ϵ*_*j*_, namely the age-specific proportion of vaccinated individuals, receives an effective vaccine. The average time required to achieve immunity from an effective vaccine is 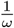.Additionally, it is assumed that after 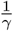, protected individuals lose immunity and become susceptible to influenza.

### Epidemiological Inputs

Vaccine efficacy varies with age, with a lower effectiveness observed in older populations [24]. Based on the literature [25, 26], we used the following vaccine effectiveness rates: 62.5% for G1, 54% for G2 and G3, and 47.8% for G4. Our model was calibrated using case data from the HIRA database in the Republic of Korea, which included the age distribution of influenza cases. We estimated the transmission coefficient for different age groups (*β*_*j*_) and seasonality constant (*α*_0_). The remaining model parameters were derived from the published literature (Table 1). Because it is difficult to estimate the value of each compartment at the beginning of an epidemic, August of 2023, which had a low and unchanged number of confirmed cases, was selected to determine these values. In equation (1), no change in compartment *X* implies *dX/dt* ≃ 0 during that period, and the confirmed cases are *C*_*j*_ = *qκE*_*j*_ for the *j*^*th*^ age group. The corresponding calculation is provided in the Supplementary Information (see S1 File).

**Table 1.**
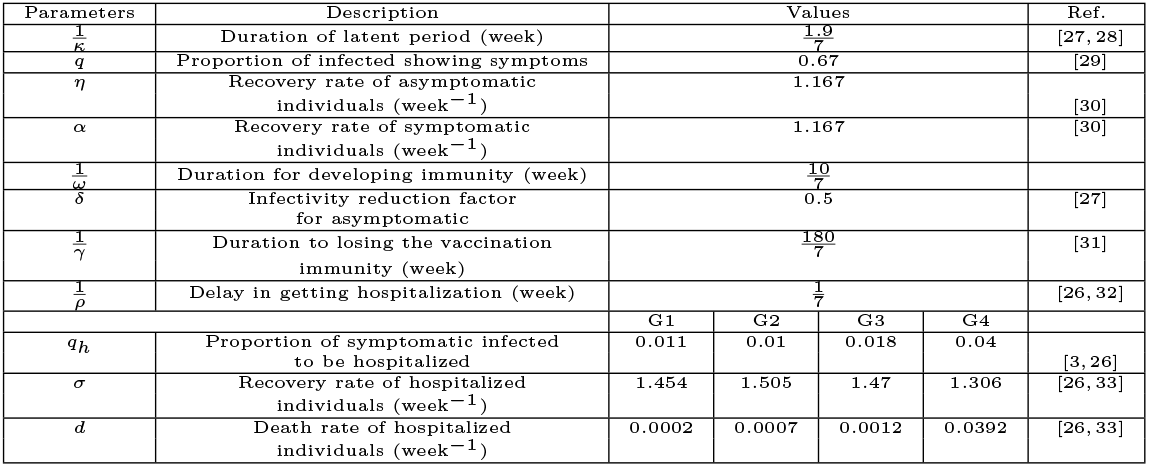
Numerical values and descriptions of model parameters.

### Vaccination scenarios with different timings

Three distinct vaccination scenarios were considered, each based on different timing strategies for administering the vaccine across various age groups. However, in all scenarios, the duration of vaccination and total number of vaccines administered remained the same for all age groups. The vaccination strategy outlined by the KNIP in the Republic of Korea was used as the baseline vaccination scenario (S0). In this baseline scenario, vaccination for the G1 group began on October 9th, and for the G4 group, it began on October 16th. Additionally, vaccination for pregnant women began on October 9th and was administered over a span of five weeks. In Vaccination Scenario-1 (S1), the timing of vaccination for each group followed the baseline strategy, with vaccinations starting on October 9th for all groups as per the KNIP guidelines. In Vaccination Scenario-2 (S2), the G4 group was prioritized, and vaccination for this group began two weeks earlier than in the baseline scenario, aiming to assess the impact of an earlier vaccination start for this particular group. The different vaccination scenarios and specific timings for the vaccination of each group are illustrated in Figure This diagram provides a visual representation of the variations in vaccination timing across the three scenarios and offers a comprehensive overview of the strategies under consideration.

## Results

### Parameter Estimation

The transmission coefficients *β*_*j*_ for different age groups were estimated by fitting the model output *qκE*_*j*_ for *j* = 1, 2, 3, 4 to the observed number of influenza cases in model (1). We used the standard *fminsearch* function in MATLAB to minimize the error between the model output and actual influenza cases. The estimated parameters were obtained as follows: *β*_1_ = 4.1604, *β*_2_ = 0.3042, *β*_3_ = 0.1071, *β*_4_ = 0.2720 and *α*_0_ = 2866. The model output fitting is depicted in Figure 3. Our parameter estimation results indicate that the transmission coefficient for G1 is the highest, followed by those for G2, G4, and G3. This pattern is consistent with the characteristics of influenza, which tends to spread more readily among younger individuals [34]. According to the HIRA data, 28.58% of individuals in G1 contracted the infection during one season, which is the highest rate among all age groups. In comparison, 7% of G2, 2.27% of G3, and 1.21% of G4 were infected during the simulation period in South Korea. Our model also produces results that align with these trends (see Figure 10 in the Supplementary Materials).

**Fig 1.**
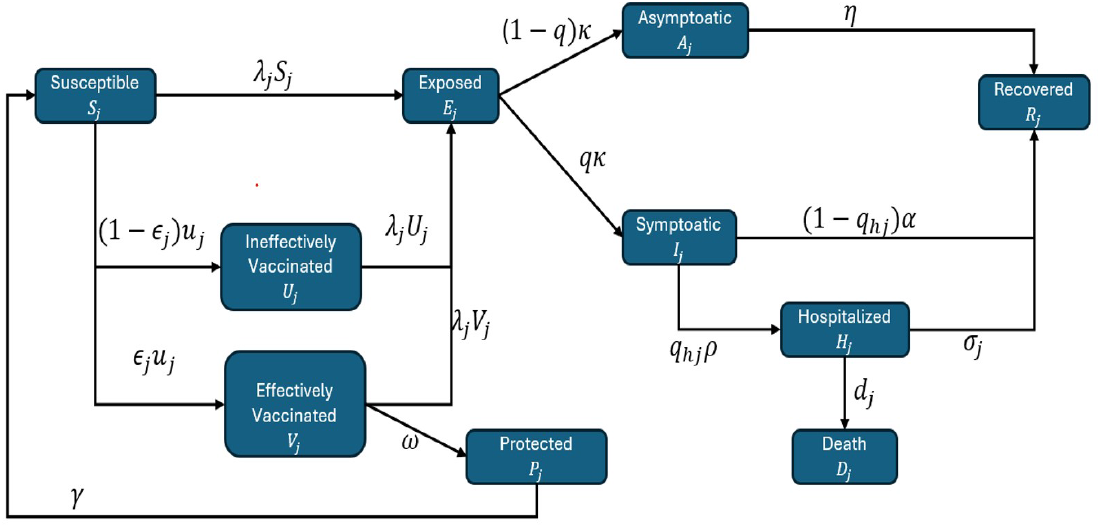
Flow diagram showing the transition of the population across different compartments.

**Fig 2.**
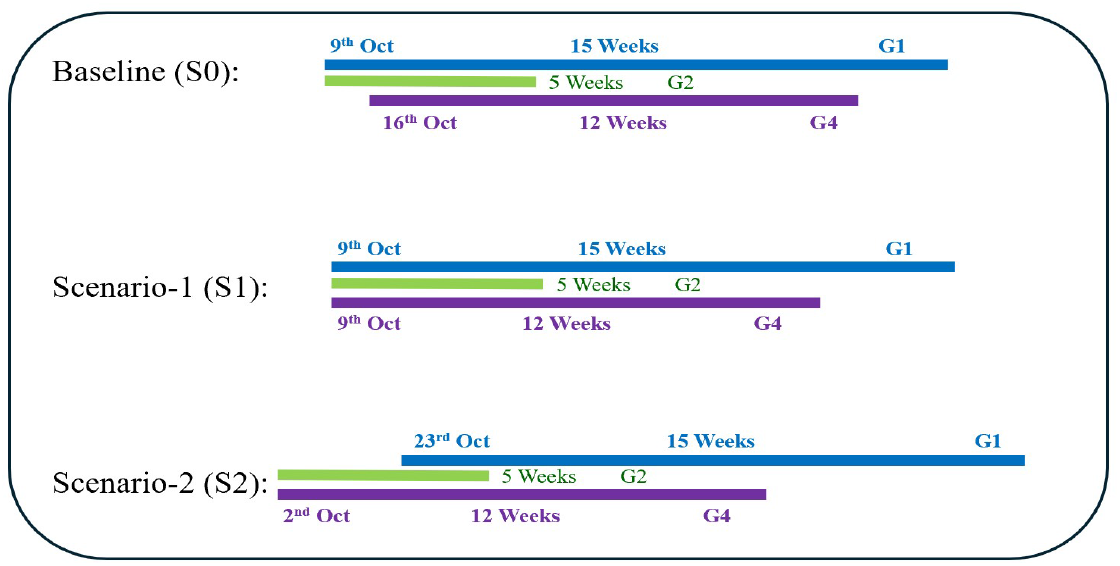
Three vaccination scenarios based on the timing of vaccination in each group.

**Fig 3.**
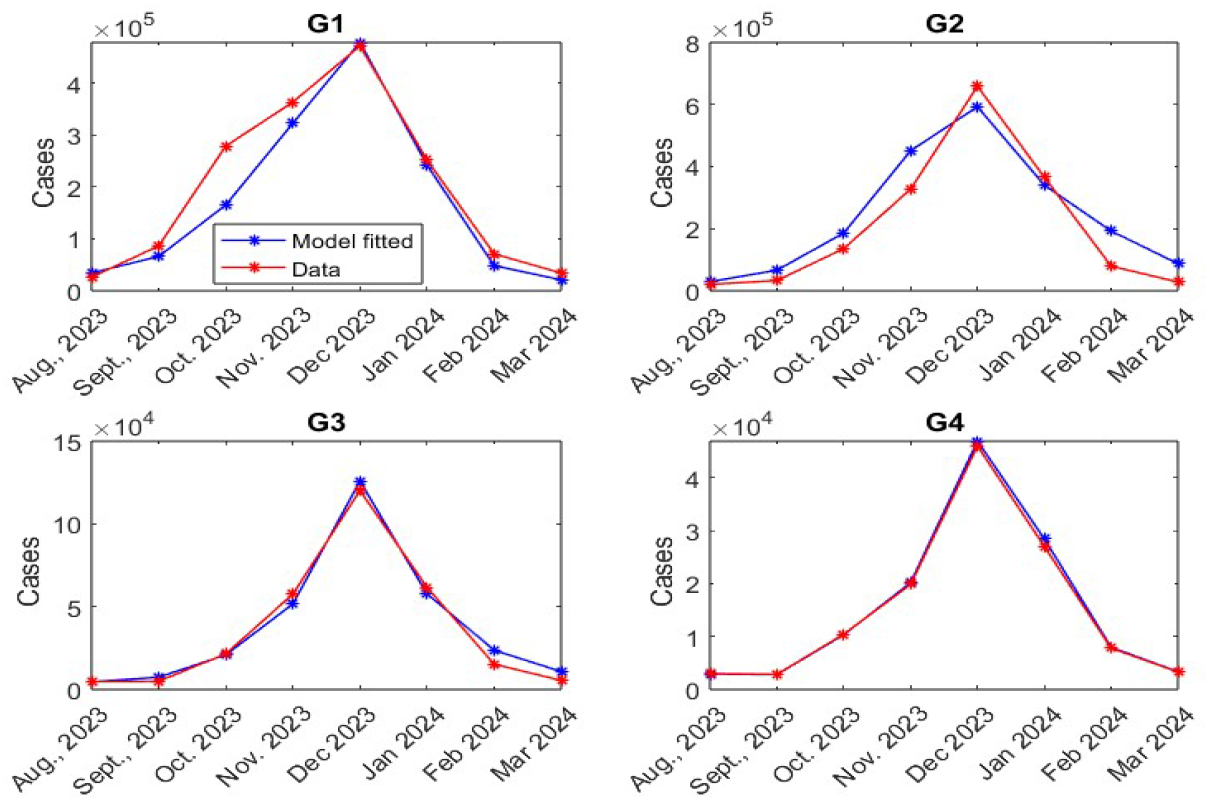
Model outputs compared with actual data, with the number of influenza cases shown as red curves and corresponding model outputs for the estimated parameters shown as blue curves.

Given the highest infection rate observed in G1, it follows that the transmission coefficient for this group is the highest, reflecting the increased susceptibility and transmission dynamics within this age group.

### Impact of Vaccination Shifting

As shown in Figure 4, we examine the impact of shifting vaccination timing on cumulative influenza cases by exploring three distinct scenarios: (1) shifting the baseline vaccination strategy (i.e., current vaccination strategy as per the KNIP), (2) shifting vaccination for the G1 group only, while keeping the vaccination strategy for the other groups unchanged, and (3) shifting vaccination for the G4 group only, while maintaining the baseline vaccination strategy for the other groups. For each of these scenarios, we analyzed the effects of shifting vaccination two weeks earlier, one week earlier, one week later, and two weeks later, and observed how these changes affected the cumulative cases across different age groups and the overall population.

**Fig 4.**
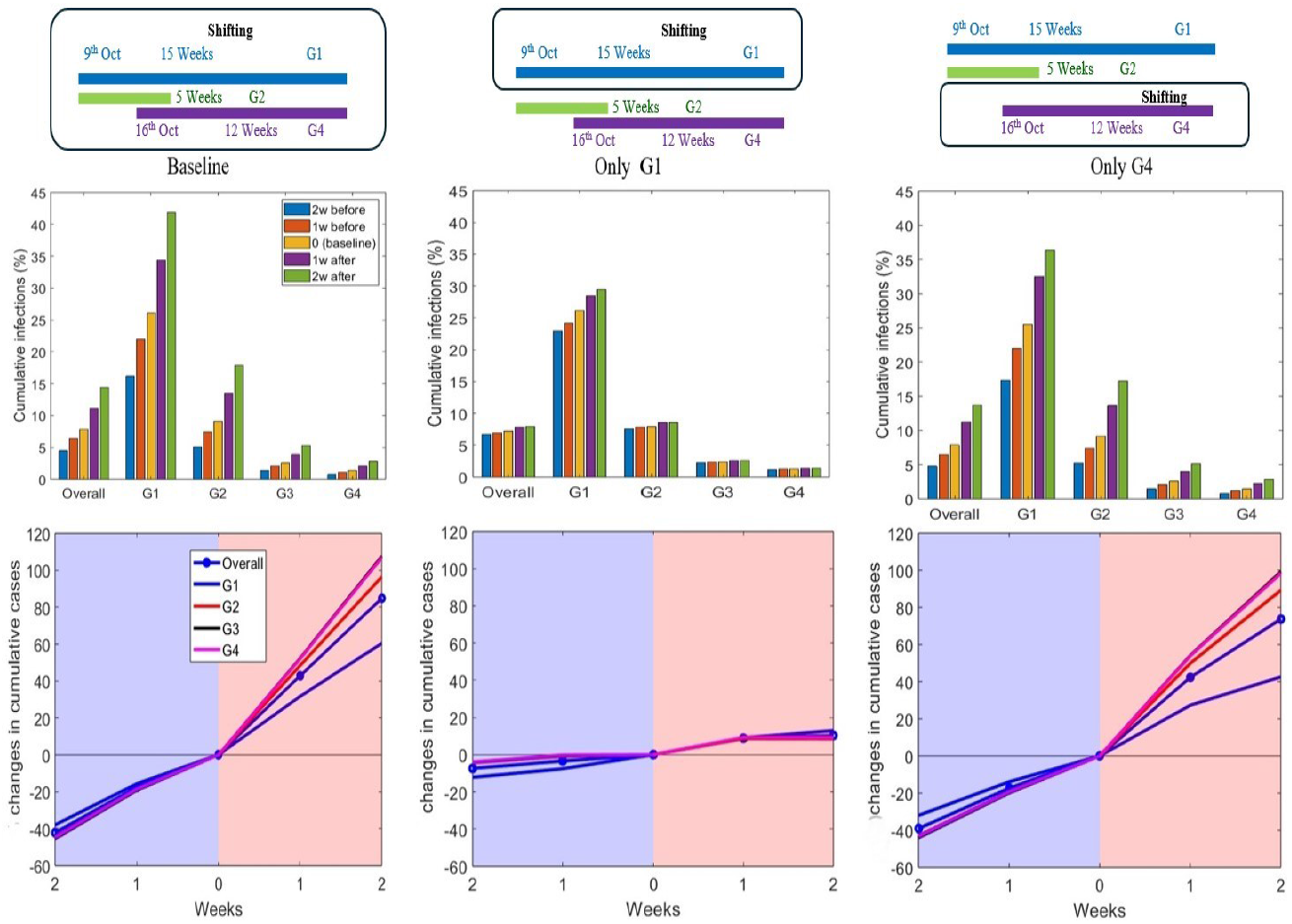
Impact of vaccination timing on cumulative cases.

When we shifted the baseline vaccination to two weeks earlier, we observe a substantial reduction in cumulative cases across all age groups, with an approximately 40% decrease in the percentage of cumulative cases for each group. However, when vaccination is delayed by two weeks, the increase in cumulative cases range from 60% to 107% across different age groups. The G4 group experiences the largest increase in cumulative cases (107%), whereas the G1 group exhibits the smallest increase (60%). The smaller increase in G1 can be attributed to the high percentage of cumulative cases in this group under current vaccination strategies. In contrast, the G4 group has the lowest percentage of cumulative cases under the current vaccination strategy, making it more vulnerable to a significant increase in cases of delayed vaccination.

When vaccination is shifted only for the G1 group, the changes in cumulative cases are relatively small. In contrast, when vaccination is shifted only for the G4 group, the observed changes in cumulative cases are similar to those observed when the baseline vaccination strategy is shifted. Specifically, the G1 group exhibits less variation in cumulative cases when the vaccination shift is applied only to the G4 group, likely because the vaccination strategy for G1 remains unchanged as per the baseline.

Therefore, the relatively smaller impact on G1 cases can be attributed to the fact that no adjustment is made for the vaccination timing of this group. A similar impact of vaccination timing on the number of deaths across different groups is observed, as shown in Figure 11 in the Supplementary Materials.

It may be concluded that the timing of vaccination for the G4 group is crucial in drastically lowering the number of seasonal influenza infections in South Korea based on the effects of vaccine timing observed across all age groups. Our results underline the significance of prompt vaccination in this age group by indicating that modifying the vaccination schedule for the G4 group, specifically by administering the vaccine earlier, significantly reduces influenza cases. Advancing the vaccination schedule for G4 can significantly reduce the overall burden of influenza and prevent the virus from spreading, as this group typically has better vaccination coverage and a comparatively lower incidence of the disease under existing policies. Therefore, prioritizing vaccination shifts for the G4 group could have a profound impact on the management of seasonal influenza outbreaks in South Korea.

### Vaccination Scenarios Based on Timing

In the left panel of Figure 5, the percentage of cumulative cases for each age group and overall cumulative cases are presented for each vaccination scenario. The right panel of Figure 5 presents the changes in the cumulative cases for scenarios S1 and S2 compared with the baseline scenario.

**Fig 5.**
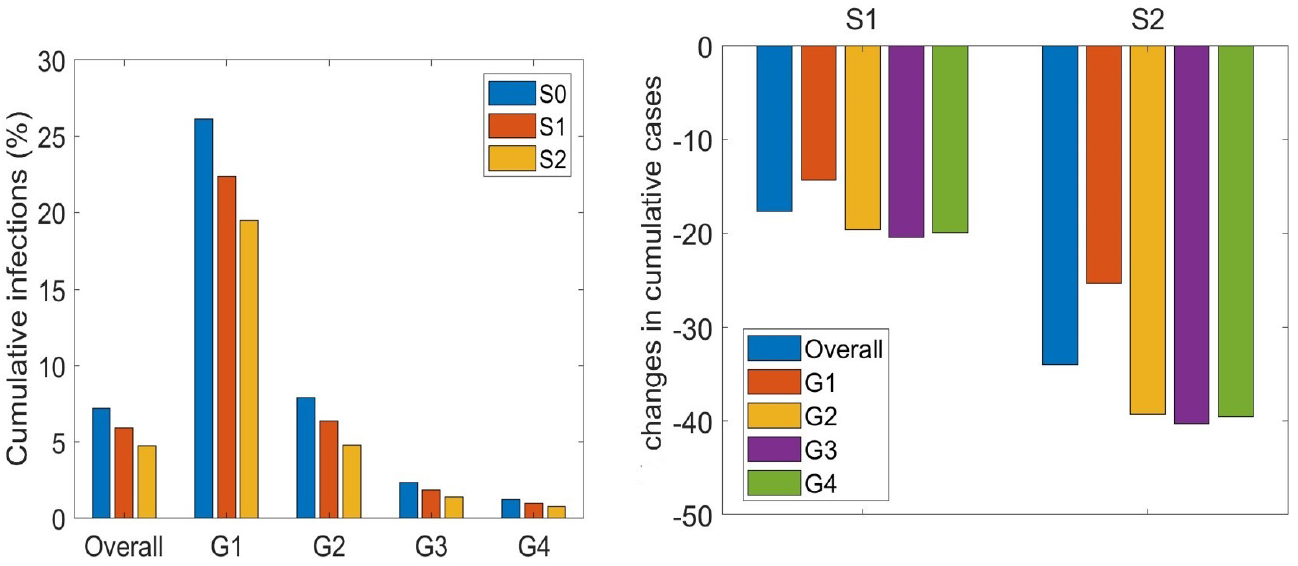
Cumulative cases for three different vaccination scenarios.

The overall cumulative cases in S1 are reduced by 17Therefore, it can be concluded that S2 would be more beneficial for reducing influenza cases in South Korea. In S2, the G4 group is prioritized and vaccination for this group begins two weeks earlier than in the baseline scenario. This prioritization of the G4 group through the earlier start of vaccination leads to a more significant reduction in cases. Therefore, prioritizing the G4 group for vaccination in South Korea could considerably reduce the number of influenza cases in the country.

### Impact of Vaccine Effectiveness, Coverage, and Timing in the G4 Group

In subsection , we observe that influenza cases are strongly influenced by vaccination in the G4 group. Subsection highlights that S2 is more effective at reducing influenza cases in South Korea. Building on these findings, we explored the impact of vaccine effectiveness, coverage, and timing, specifically for the G4 group, within S2. In S2, the vaccine coverage and effectiveness for the G4 group were 82.5% and 47.8%, respectively. The left panel of Figure 6 reveals that even with vaccine coverage lower than that in S2, influenza cases can still be reduced by increasing vaccine effectiveness in the G4 group. This finding suggests that improving vaccine effectiveness may compensate for lower vaccine coverage. A similar pattern is observed in the right panel of Figure 6, where the reduction in influenza deaths follows a similar trend, with an increasing vaccine effectiveness leading to a decrease in the number of deaths, even with lower coverage. To increase vaccine effectiveness, alternative vaccines such as the adjuvanted quadrivalent influenza vaccine should be considered. This vaccine has higher efficacy than the standard quadrivalent influenza vaccine [25, 35] and its use in the G4 group could further enhance the impact of vaccination. By improving both vaccine coverage and effectiveness, particularly with alternative vaccines, the reduction in influenza cases and deaths could be further optimized in the G4 group, thereby contributing to improved disease control in the Republic of Korea.

**Fig 6.**
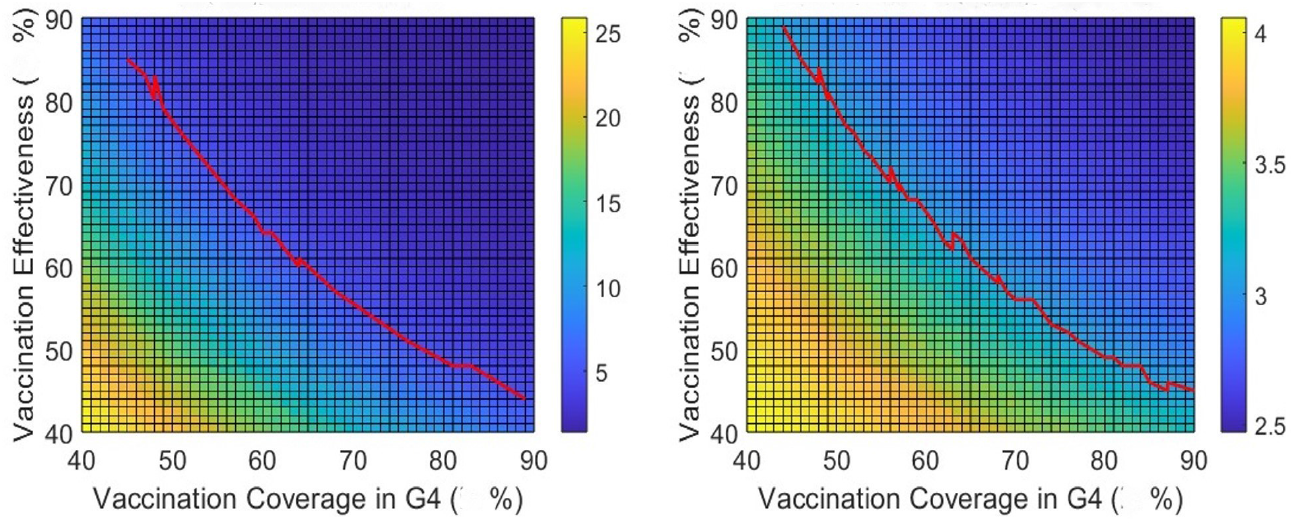
Cumulative cases percentage (left panel) and deaths in log10 scale (right panel) for different vaccine effectiveness and coverage values in the G4 group. The red curve represents the percentage of overall cumulative cases and cumulative deaths, which are identical to those observed in S2.

As shown in the left panel of Figure 7, even with vaccine coverage lower than that in S2, influenza cases could still be reduced by administering vaccination earlier in the G4 group. Specifically, by providing vaccination two weeks earlier than in S2, the cumulative number of influenza cases are reduced, even at a vaccination coverage of 75%. This result demonstrates that advancing the vaccination timeline for the G4 group can lead to a significant reduction in the number of cases, even when the coverage is lower than that in S2.

**Fig 7.**
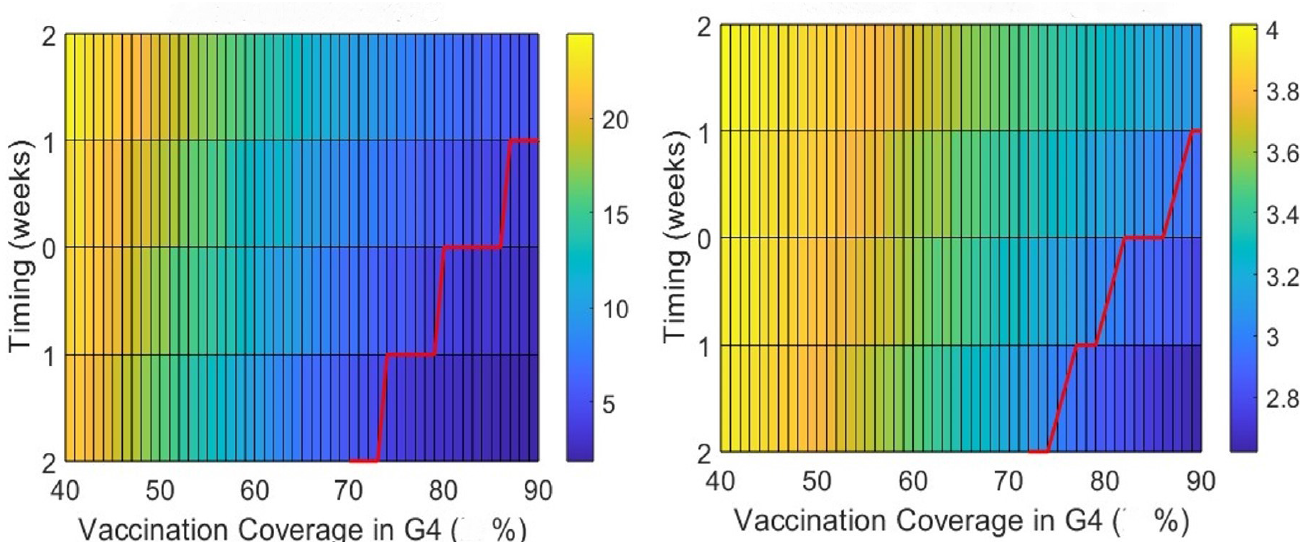
Cumulative cases percentage (left panel) and deaths in log10 scale (right panel) for different vaccine timings and coverages in the G4 group . The red curve represents the percentage of overall cumulative cases and cumulative deaths, which are identical to those observed in the S2 scenario.

A similar trend is evident in the right panel of Figure 7, where influenza-related deaths follow a comparable pattern. By shifting the vaccination timing for the G4 group to an earlier start, influenza-related deaths are also reduced, mirroring the trend observed in the cumulative cases. These results emphasize the importance of vaccination timing in reducing both influenza cases and deaths, particularly when earlier vaccination is implemented in the G4 group.

### Impact of natural immunity on the peak

In this study, we considered three distinct levels of immunity: (1) L1: Immunity resulting from the reported cases of the previous year; (2) L2: Immunity resulting from both reported and unreported cases of the previous year; and (3) L3: Immunity from reported and unreported cases of the previous year, along with 50% of the cases from the previous year. Figure 8 illustrates the peak size of the cumulative influenza cases under different vaccination scenarios and immunity levels. As expected, it is evident that the model predicts the smallest peak in S2 when the L3 immunity level is applied. This suggests that a higher level of immunity, which accounts for both reported and unreported cases from the previous year, as well as a portion of cases from the previous year, leads to a substantial reduction in the peak of influenza cases in S2. Table 2 presents the timing of the peak of the cumulative cases for each vaccination scenario and immunity level. In S0 and S1, the peak of cumulative cases occurs with a delay of one to two weeks as the immunity level increases. This implies that higher immunity levels slow the onset of the peak, possibly as a result of reduced susceptibility to infection within the population. In contrast, for S2, no changes are observed in the timing of the peak, regardless of the immunity level, suggesting that the timing of the peak is unaffected by the varying immunity levels in this vaccination strategy.

**Table 2.**
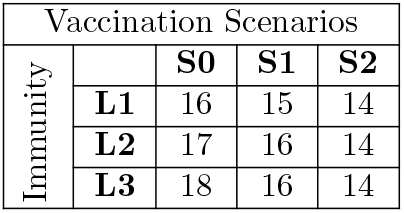
Timing of the peak for different vaccination scenarios and immunity levels (in which week the peak of cases is attained).

**Fig 8.**
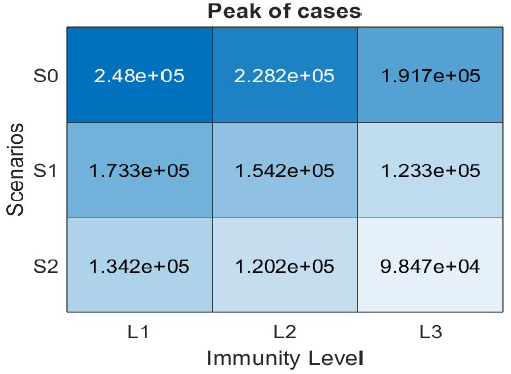
The peak size for different vaccination scenarios and immunity levels.

In our analysis, we examined two distinct thresholds for the number of hospitalized individuals. Threshold-1 was set at 4000 and threshold-2 at 5000. The goal of this analysis was to assess the hospital burden resulting from seasonal influenza. Figure 9 presents the duration over which the number of hospitalized individuals surpassed each threshold. This figure reveals how the burden on hospitals varies under different vaccination scenarios and immunity levels. Figure 9 and Table 3 indicate that the duration for which the number of hospitalized individuals exceeds the threshold is the longest in S0, which represents the baseline scenario. In contrast, S2 consistently results in a shorter duration of hospital burden, exceeding both thresholds across all immunity levels considered in the study. This suggests that S2, which likely involves a more effective or widespread vaccination strategy, is the most impactful in terms of reducing the strain on hospitals due to influenza compared with the other vaccination scenarios explored in our analysis. Therefore, we conclude that S2 offers the most significant potential for mitigating the hospital burden of seasonal influenza, thereby demonstrating its efficacy in controlling the impact of the disease on healthcare systems.

**Table 3.** Duration (in weeks) for which the number of hospitalized individuals exceeds the threshold.

| Vaccination Scenarios |  |  |  |  |
| --- | --- | --- | --- | --- |
| Immunity |  | S0 | S1 | S2 |
|  | L1 | 17 | 17 | 14 |
|  | L2 | 18 | 16 | 11 |
|  | L3 | 18 | 14 | 4 |
| Immunity |  | S0 | S1 | S2 |
|  | L1 | 16 | 13 | 7 |
|  | L2 | 16 | 13 | 2 |
|  | L3 | 15 | 9 | 0 |

(a) When hospitalized threshold = 4000
(b) When hospitalized threshold = 5000

**Fig 9.**
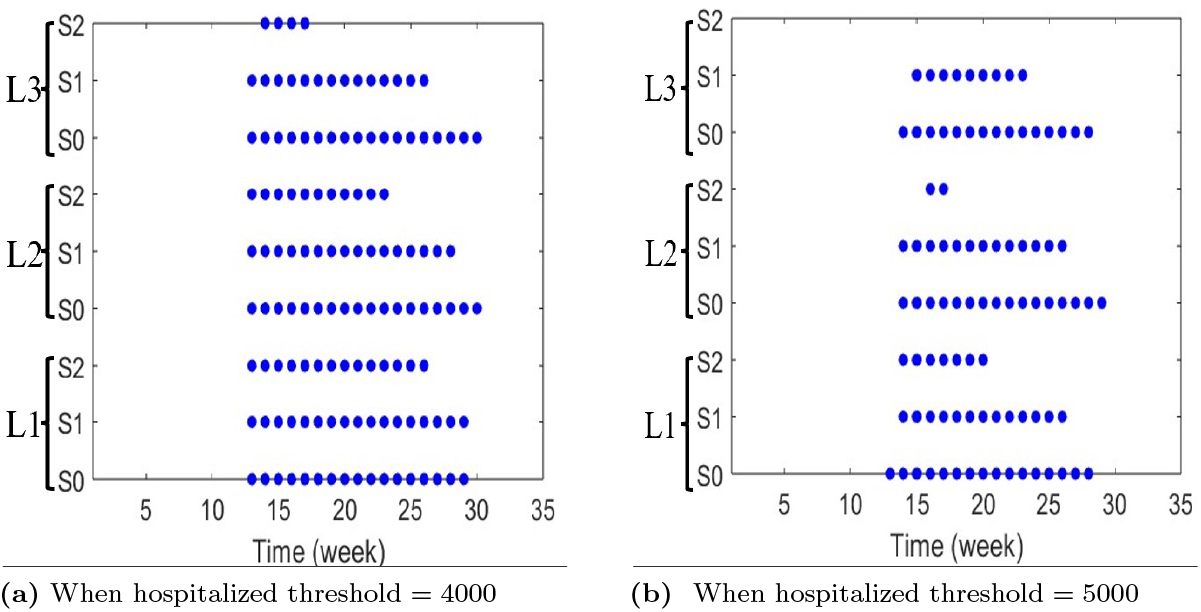
Time during which the number of hospitalized individuals exceeds the threshold

**Fig 10.**
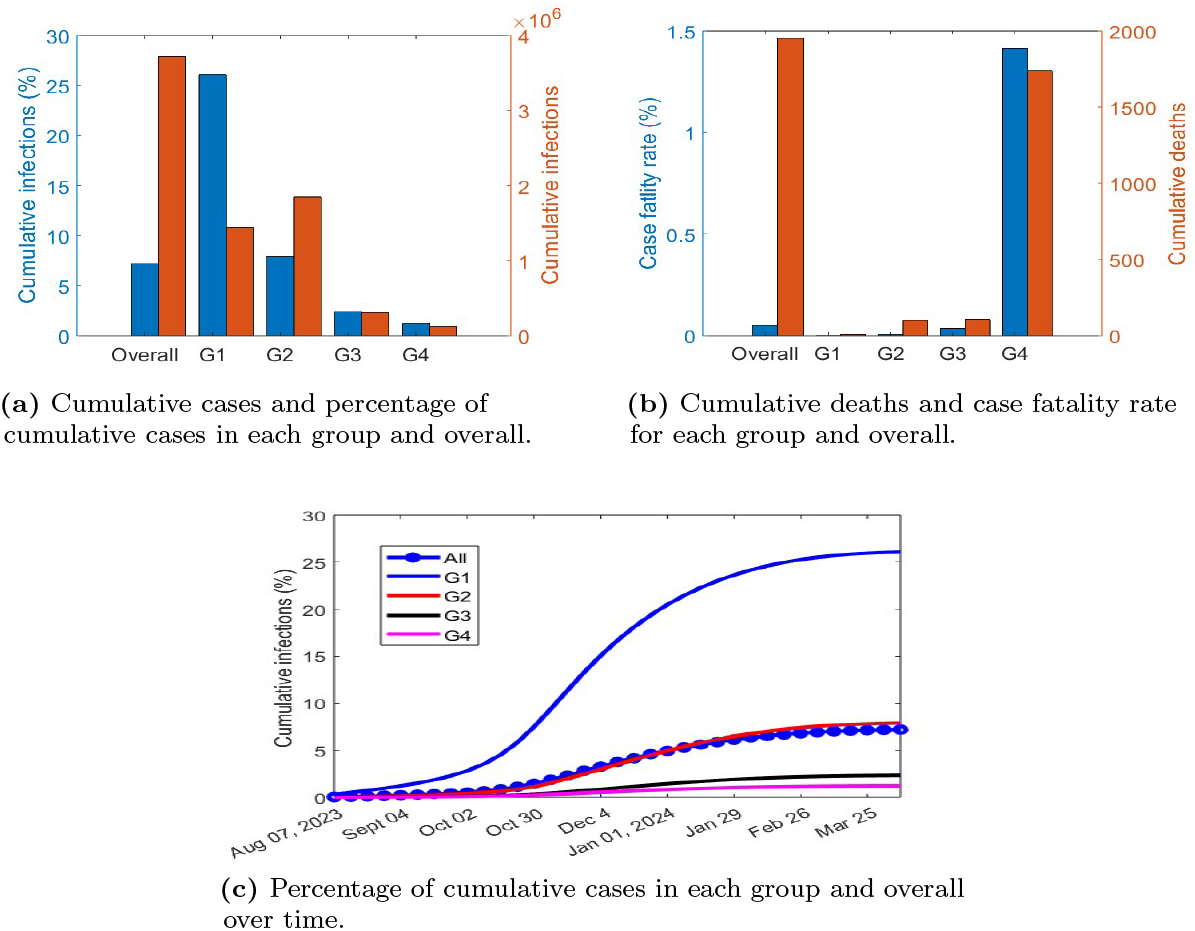
Model outputs of estimated parameters (*β*_*j*_ and *α*_0_)

**Fig 11.**
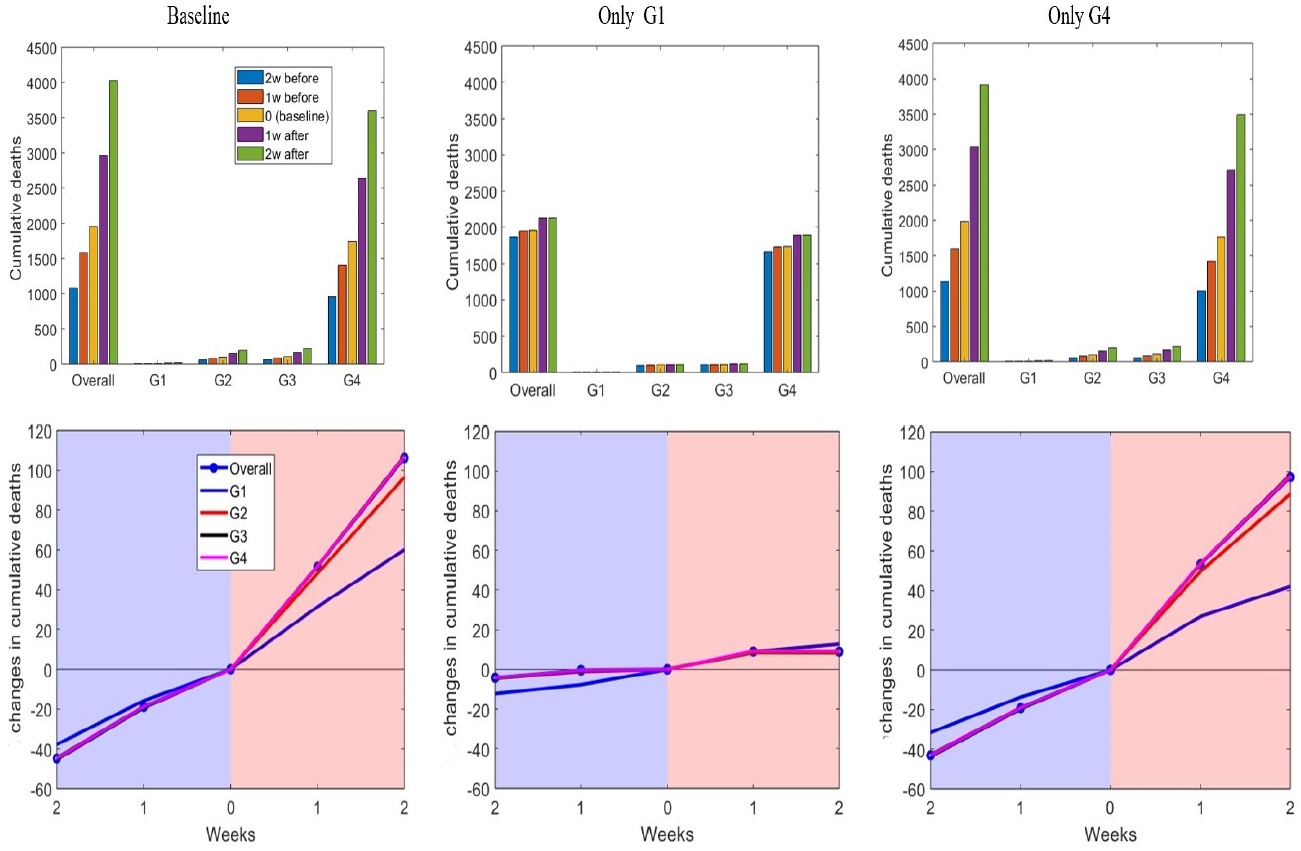
Impact of vaccination timing on cumulative deaths of each group and overall.

**Fig 12.**
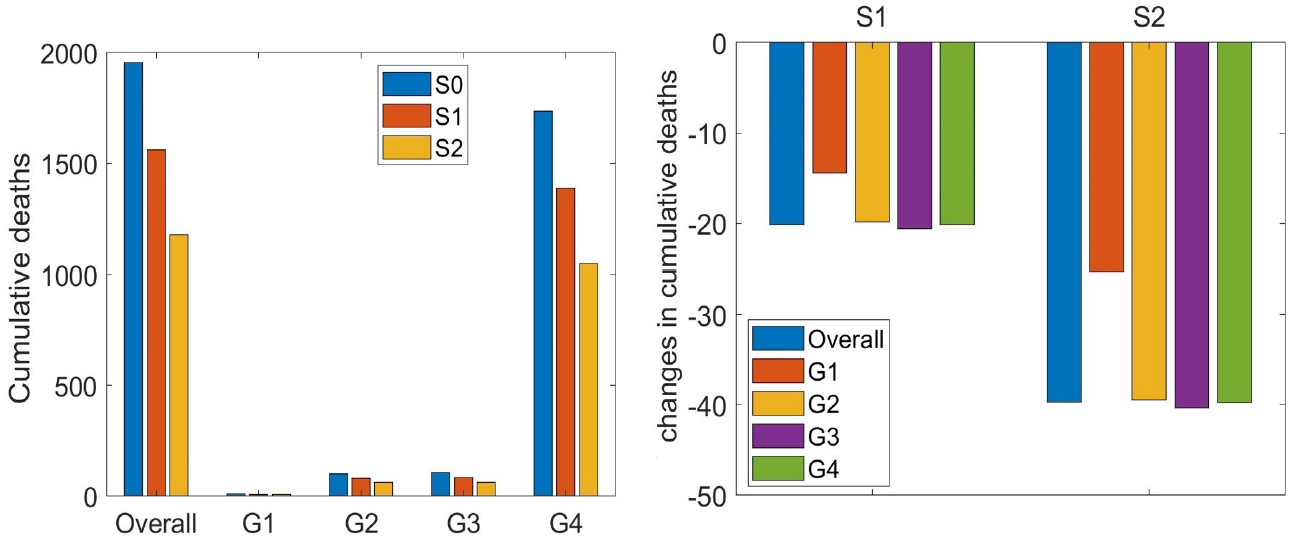
Cumulative deaths for three different vaccination scenarios and changes in cumulative deaths for S1 and S2 compared with S0 (baseline).

**Fig 13.**
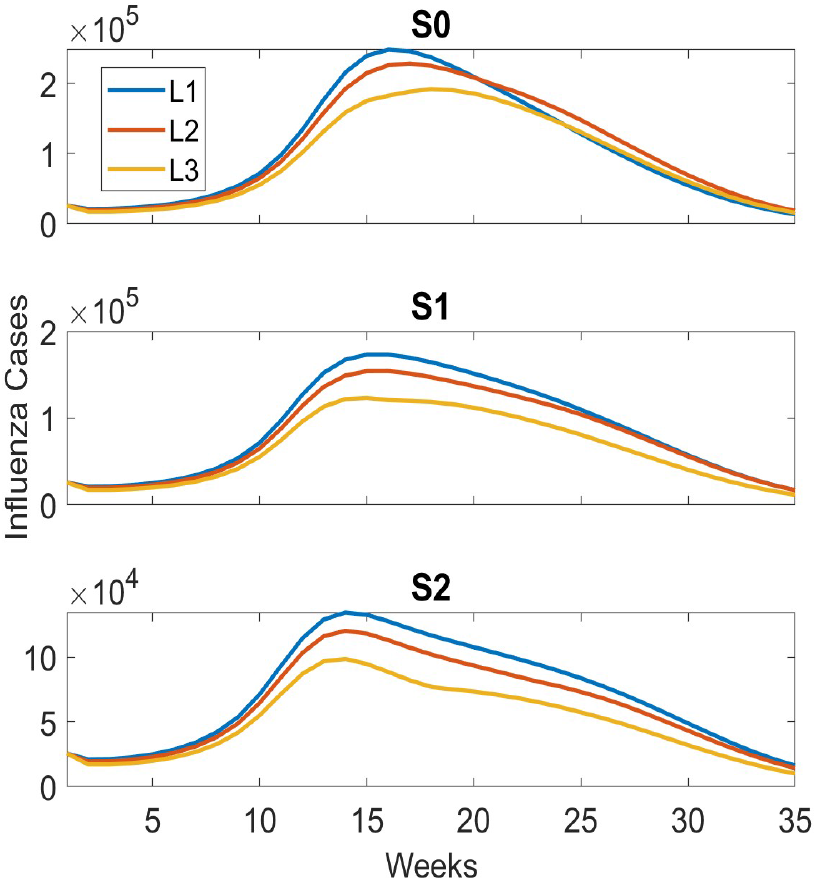
Cumulative cases for three different vaccination scenarios and immunity levels over time.

**Fig 14.**
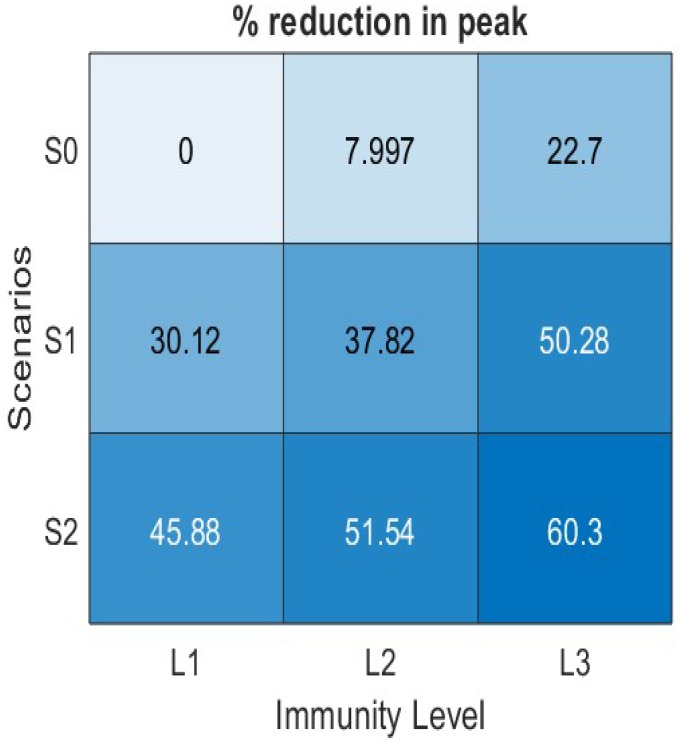
Percentage reduction in the peak size of cumulative cases for different vaccination and immunity levels. Here, S0-L1 is considered the baseline case.

**Fig 15.**
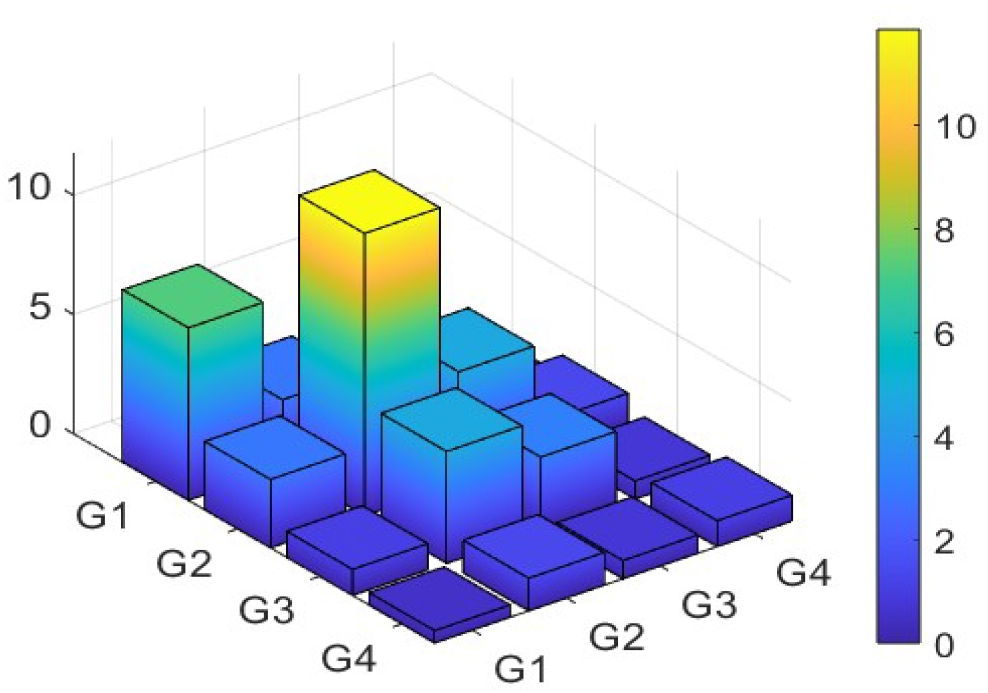
Contact matrix for four age groups.

**Fig 16.**
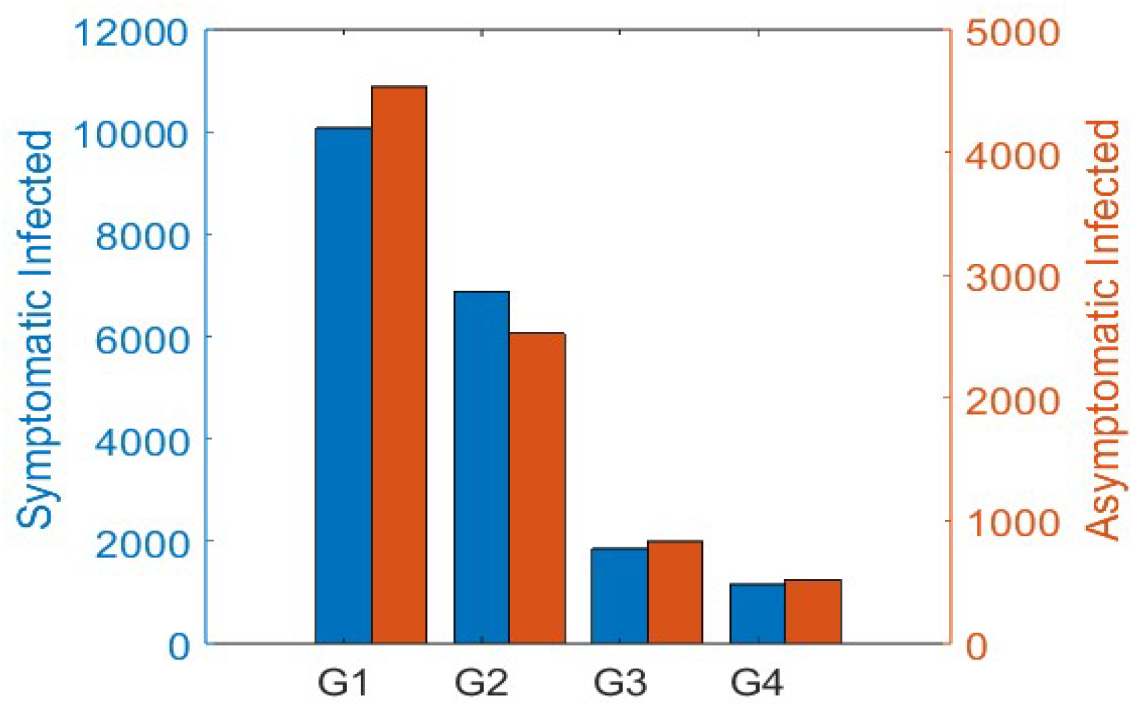
Initial infected population for each age group.

## Discussion

The potential for a devastating influenza pandemic characterized by high rates of morbidity and mortality, particularly among the elderly, presents a significant challenge to global public health. In situations where pharmaceutical tools are scarce or unavailable, controlling the spread of the disease relies heavily on non-pharmaceutical interventions such as the use of facial masks, social distancing, travel restrictions, and isolation of infected individuals. The implementation of such measures could provide valuable time, allowing the mass production of antiviral medications and vaccines to meet the needs of large populations [36–38]. In addition to these non-pharmaceutical strategies, policies aimed at reducing the impact of seasonal influenza must consider pharmacological measures that can mitigate the severity of the epidemic. The efficient prioritization of vaccine distribution is a critical component of these efforts, requiring careful consideration of several factors, including population demographics, levels of background immunity, and the availability of resources [39, 40].

In this study, we developed a mathematical model to assess the transmission dynamics of seasonal influenza considering age-related differences in disease susceptibility, transmission rates, hospitalization, and mortality. Our model incorporated time-dependent vaccination rates, allowing us to explore different vaccination strategies under varying conditions of immunity and vaccination timing.

Our results highlight that the highest infection rates were observed in the G1 group (children aged 0 to 14 years), which is consistent with the increased susceptibility and heightened transmission dynamics within this age cohort. As a result of their high levels of social interaction and activity, this group contributes most significantly to the overall spread of influenza. Under the baseline vaccination strategy, which aligns with the current KNIP in the Republic of Korea, 25.16% of individuals in the G1 group, 7.90% in the G2 group (15 to 49 years), 2.34% in the G3 group (50 to 64 years), and 1.23% in the G4 group (65 years and older) were infected during the 2023 to 2024 flu season. These findings are consistent with the data from HIRA [18, 19]. Additionally, we examined the impact of vaccination timing on various age groups and found that vaccinating the G4 group earlier in the season significantly reduced the number of infections, hospitalizations, and deaths. Notably, delayed vaccination in the G4 group diminishes the overall effectiveness of the vaccination campaign.

Our analysis further reveals that prioritizing vaccination in the G4 group (those aged 65 years and older) could substantially reduce the burden of the epidemic. In scenarios where vaccination is prioritized for this age group, the cumulative cases decrease by approximately 34% (S2) and 17% (S1) compared with the baseline scenario. Advancing the vaccination schedule for the G4 group leads to a marked reduction in the peak number of cases, as individuals in this high-risk group contribute significantly to the severity of the epidemic. Even in scenarios with lower vaccine coverage than S2, improving the effectiveness of the vaccine for the G4 group could lead to a notable reduction in influenza cases. This finding suggests that enhancing vaccine efficacy, such as using adjuvants or alternative quadrivalent vaccines, could partially compensate for the lower vaccine coverage in this critical age group [25, 26].

In terms of practical outcomes, the S2 vaccination strategy prioritizing the G4 age group reduces the peak number of cases by 45% to 60% depending on varying levels of immunity. Furthermore, it shortens the duration required for the pandemic to surpass different hospitalization thresholds. These results underscore the importance of early and targeted vaccination efforts, particularly for high-risk groups, mitigating the public health impact of seasonal influenza. The WHO has established guidelines for prioritizing immunization based on the epidemiology of influenza viruses, as well as morbidity, mortality, and vaccine efficacy data. These guidelines are designed to minimize the number of infections, hospitalizations, and deaths, while addressing the broader socioeconomic impacts of an influenza pandemic. Although developed countries typically secure and distribute sufficient vaccine supplies, the availability of vaccines in developing nations during outbreaks remains a critical concern. Therefore, it is crucial not only to increase global vaccine production but also ensure equitable distribution, particularly in countries with limited resources.

Future research should consider asymptomatic cases, other high-risk populations, and the constraints imposed by current vaccine technologies, including delays in vaccine deployment following the onset of a pandemic. Our findings also indicate the potential benefits of pre-pandemic immunization with vaccines tailored to pandemic strains, although further studies are required to assess the long-term effects and safety of such strategies [41].

Overall, this study contributes valuable insights into the design of age-specific, optimal vaccination strategies that can effectively reduce influenza incidence, aimed at reducing the incidence of influenza, as well as the associated hospitalizations and mortality rates. By improving our understanding of how vaccination can be optimized across different age groups, our findings can inform policymakers in implementing timely and effective responses to influenza outbreaks, ultimately reducing both the health and economic burdens of these seasonal outbreaks. These strategies can help ensure that high-priority populations such as the elderly are effectively protected, even in the context of limited resources. Furthermore, identifying the optimal timing and distribution of vaccines across various age groups is crucial for the development of future vaccine delivery plans, particularly in the face of the rapidly evolving influenza threat. As we did not consider the waning of immunity due to recovery, it was difficult to investigate long-term predictions based on vaccination strategies. To consider waning immunity related to recovery, antibodies and genotypes should be considered simultaneously [42]. Every year, the genotype of influenza has changed and vaccine companies have invented new vaccines considering new genotypes that are expected to spread in each year.

## Data Availability

All relevant data are within the manuscript and its Supporting Information files.

## Acknowledgments

This paper is supported by the Korea National Research Foundation (NRF) grant funded by the Korean government (MEST) (NRF-2021R1A2C100448711). This research is also supported by the Bio&Medical Technology Development Program of the National Research Foundation (NRF) funded by the Korean government (MSIT) (RS-2023-00227944). The funders had no role in study design, data collection and analysis, decision to publish, or preparation of the manuscript.

## Supporting information

**S1 Fig. Model Outputs of Estimated Parameters**

**S2 Fig. Impact of vaccination timing on cumulative deaths**.

**S3 Fig. Vaccination scenarios**

**S4 Fig. Vaccination scenarios and immunity levels**.

**S5 Fig. Reduction in the peak size of cumulative cases**

**S1 Table. Duration (in weeks) for which the number of hospitalized individuals exceeds the threshold**

**S1 File. Supplementary Material**

### Contact Matrix

The estimated contact matrix for the Republic of Korea presented by Prem et al. [43] was derived using population-based contact diaries from the European POLYMOD survey. The survey was used to generate projections for various countries, including the Republic of Korea. These contact matrices contain contact rates across 16 age categories organized in five-year increments, covering the age range of 0 to 80 years.

However, for the purpose of our model, we focused on only four broad age groups. To this end, we aggregated the original data into four distinct age groups: 0 to 14, 15 to 49, 50 to 64, and 65 to 80 years old. This aggregation was performed by calculating the population-weighted sums across individual age classes within each broader group. The following population-weighted sum was used:

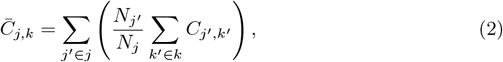

where *{j*^*′*^, *k*^*′*^*}* are the subscripts for the five-year age intervals, *{j, k}* are the subscripts for the larger age intervals we considered, *C*_*j*_^*′*^,*k*^*′*^ represents the average number of daily contacts a person in age group *j*^*′*^ makes with group *k*^*′*^, and *N*_*j*_^*′*^ is the total population of age group *j*^*′*^.

The total number of *j*–*k* contacts must be equal to that of *k*–*j* as *N*_*j*_*C*_*j*,*k*_ = *N*_*k*_*C*_*k*,*j*_. As a result of the numerical challenges encountered during estimation, including bin discretization and rounding errors, small discrepancies can arise in the results. To address this issue, we took precautionary measures to ensure that the conditions were consistently met by implementing specific conditions. This approach helps minimize the impact of such issues and ensures that the desired outcome is achieved. We ensured that this condition holds true by imposing

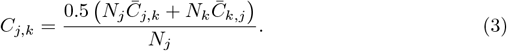

Here, the numerator represents the average of the two measures of the total contact between groups *j* and *k*, whereas the denominator adjusts the result to a per-capita basis for group *j*. From this process, we obtained the contact matrix shown in Figure 15.

### Initial Conditions

To estimate the initial size of the various model compartments, we utilized the monthly confirmed influenza case data from the HIRA [18, 19] and weekly data from the FlueNet surveillance system of the WHO [20]. When comparing the two datasets, we observed similar patterns. This approach allowed us to distribute the monthly HIRA data at weekly intervals to derive the confirmed case data for the first week of August of 2023.

From the confirmed case data for the first week of August of 2023, we estimated the number of individuals exposed at the initial time *t* = 0 (i.e., first week of August). The number of exposed individuals was calculated based on the confirmed cases at *t* = 0 using the following relationship:

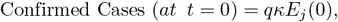

where*j* = 1, 2, 3, 4 denotes to the different age groups. To refine the model further, we assumed that the confirmed cases observed in the first week of August of 2023 were indicative of a stable state. Therefore, we assumed that the rates of change in the infected compartment, namely 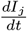 and 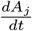,were zero at *t* = 0. This assumption allowed us to calculate the initial population sizes of infected (*I*_*j*_) and asymptomatic (*A*_*j*_) individuals for each age group at *t* = 0.

To calculate the recovered population, we assumed that individuals who had been confirmed to be infected in the previous year were likely to have developed immunity to influenza in the current year. Therefore, confirmed cases from the previous year were considered as the recovered population for the current model.

Regarding vaccination-related compartments, we assumed that no vaccinations were administered in August of 2023. Therefore, these compartments were considered to have a population size of zero. Finally, the remaining individuals in each age group who had not been classified as exposed, infected, or recovered were considered susceptible within their respective age groups.

